# Assessing Precision Antisense Oligonucleotide Therapy Eligibility for Infantile Genetic Epilepsies

**DOI:** 10.64898/2025.12.02.25341084

**Authors:** Emma Sherrill, David Cheerie, Cara J. Beck, Ella F. Whittle, Yasin Shafi, Natalie J. Chandler, John Christodoulou, Jerusalem Daniel, Jane Hassell, Maria Lachgar-Ruiz, Sarah Mulhern, Elizabeth Scotchman, Jashanpreet Sidhu, Celine Florentia Tedja, Gene-STEPS Study Group, Lyn S. Chitty, J. Helen Cross, Ingrid E. Scheffer, Haiyan Zhou, Timothy W. Yu, Vann Chau, Sarah E. M. Stephenson, Annapurna Poduri, Katherine B. Howell, Amy McTague, Gregory Costain, Alissa M. D’Gama

## Abstract

**Importance:** The highest incidence of pediatric epilepsy is in the first year of life. Most infantile epilepsies have presumed genetic etiologies and timely precision genetic diagnosis is increasingly possible. However, treatment remains largely symptomatic and outcomes poor due to a gap from precision diagnoses to precision therapies. Antisense oligonucleotide therapies are a precision therapy approach that holds promise for transforming outcomes.

**Objective:** To determine the proportion of infants with genetic epilepsies eligible for precision antisense oligonucleotide therapy approaches.

**Design:** This cohort study assessed eligibility for antisense oligonucleotide therapy approaches for 160 infants with epilepsy enrolled in the Gene-STEPS study from September 2021 to March 2025 with diagnostic genome sequencing. Clinical data were collected through October 2025.

**Setting:** Four pediatric referral centers.

**Participants:** Participants with infantile epilepsy and diagnostic genome sequencing.

**Exposure(s):** Assessment for antisense oligonucleotide therapy eligibility using established guidelines and multidisciplinary review.

**Main Outcome(s) and Measure(s):** Primary: proportion eligible for antisense oligonucleotide therapy approaches based on variant assessment; Secondary: proportion remaining eligible after considering general disease factors and patient-specific phenotypes.

**Results:** We assessed 160 infants with genetic epilepsies (86 male (54%)), 39 with neonatal seizure onset (<44 weeks postmenstrual age, 24%)), for eligibility for precision antisense oligonucleotide therapy approaches. Of 152 unique variants, 133 were single nucleotide variants or small insertions-deletions (74 missense, 23 frameshift, 26 nonsense, 8 intronic, 2 in-frame indels), 17 copy number variants (4 intragenic), and 2 repeat expansions. Twenty-four unique variants from 25 infants (15.6%) were in principle eligible for an exon-skipping, knockdown, splice correction, or existing upregulation antisense oligonucleotide therapy approach. Taking into account general disease factors and patient-specific phenotypes, 16/25 infants (64%) could be currently considered for these approaches and an additional 5/25 (20%) could have been considered at seizure onset.

**Conclusions and Relevance:** A substantial proportion of infants with genetic epilepsies may be eligible for precision antisense oligonucleotide therapy approaches. Our findings highlight the potential of these emerging therapies to narrow the gap from precision diagnoses to precision therapies for this population.

**Key Points:** *Question:* What proportion of infants with genetic epilepsies are eligible for precision antisense oligonucleotide therapy approaches?

*Findings:* In this cohort study of 160 infants with genetic epilepsies, 25/160 infants (15.6%) had variants that are, in principle, eligible for antisense oligonucleotide therapy approaches. Taking into account both general disease factors and patient-specific phenotypes, 16/25 infants (64%) could be currently considered for these approaches and an additional 5/25 (20%) could have been considered at seizure onset.

*Meaning:* A substantial proportion of infantile genetic epilepsies may be eligible for precision antisense oligonucleotide therapy approaches.

## INTRODUCTION

Infantile-onset epilepsies, which affect approximately 1 in 1200 infants, range in severity from self-limited syndromes to severe developmental and epileptic encephalopathies (DEEs) associated with substantial morbidity and mortality.^1, 2^ Although most have presumed genetic etiologies and precision genetic diagnosis is increasingly possible, treatment remains largely symptomatic and outcomes poor due to a gap from precision diagnoses to precision therapies.^3, 4^

Proof-of-concept exists for emerging precision antisense oligonucleotide (ASO) therapies, nucleic acid-based therapies that bind endogenous RNA and are customized for a specific genetic condition or even specific genotype, in the treatment of genetic epilepsies (e.g. *MFSD8*, *SCN1A, SCN2A*, *UBE3A*).^5–9^ However, their potential utility across genetic epilepsies is unknown.

Gene-STEPS (Shortening Time of Evaluation in Pediatric epilepsy Services) is an international multicenter study that enrolls infants with unexplained new-onset epilepsy and performs rapid genome sequencing (GS) to enable timely genetic diagnosis.^10^ We aimed to determine the proportion of infants with genetic epilepsies eligible for precision ASO therapy approaches by applying two established guidelines for ASO assessment to infants with genetic epilepsies in the Gene-STEPS cohort.^11, 12^

## METHODS

### Cohort

Gene-STEPS is a flagship study of the International Precision Child Health Partnership (IPCHiP).^3^ From September 2021 to March 2025, we recruited infants with unexplained new-onset epilepsy or complex febrile seizures and performed rapid GS as previously described.^10^ We assessed 40 genetically diagnosed infants from each of the four participating sites (**eMethods**). Demographic and clinical data were collected from the electronic medical record from enrollment through October 2025 (**eMethods**).

### Eligibility Assessment

Diagnostic variant(s) for each infant were independently reviewed by two assessors using the N=1 Collaborative (N1C) VARIANT Guidelines, and also assessed for splice correction amenability using principles from Kim et al., 2023 (a slightly more permissive splice-switching framework compared to the conservative-by-design N1C guidelines), with discrepancies between assessors resolved by multi-site review.^11, 12^ Infants with variants classified as “eligible” or “likely eligible” by N1C Guidelines or “probably” or “possibly” amenable by the splice-switching framework were considered in principle eligible for ASO therapy approaches. These infants were further evaluated by multi-site review taking into account general disease factors and patient-specific phenotypes^13^ (**Figure 1**, **eMethods**).

**Figure 1:**
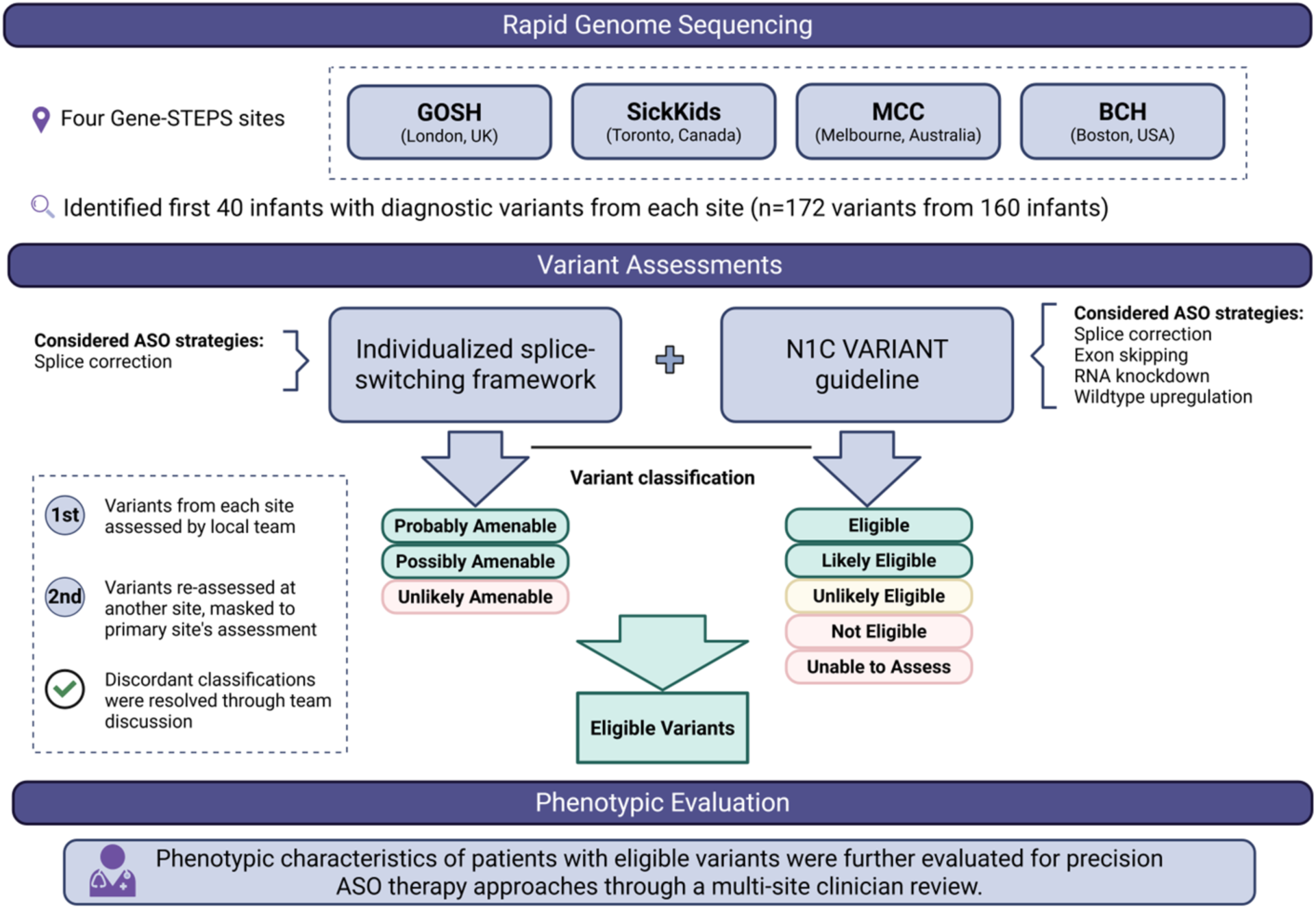
Study workflow. Diagnostic variants identified by rapid genome sequencing for infants in the Gene-STEPS cohort were assessed for ASO eligibility by two independent assessors using two established guidelines to identify in principle eligible variants (highlighted in green). Infants with eligible variants were further evaluated for general disease factors and patient-specific phenotypes through multi-site clinician review.

### Statistics

The primary outcome was the percentage of infants in principle eligible for ASO therapy approaches based on variant assessment. The secondary outcome was the percentage of the above infants who could be considered for ASO therapy approaches currently or at seizure onset, taking into account general disease factors and patient-specific phenotypes.

### Ethics

Gene-STEPS was approved by the Institutional Review Boards of participating sites. Parents provided written informed consent. Data are reported per STROBE guidelines.

## RESULTS

We assessed 160 infants with genetic epilepsies for eligibility for precision ASO therapy approaches, including 86 males (54%), 39 with neonatal-onset seizures (24%), and 79 (49%) with DEEs at onset (**Table 1**). Of the 172 diagnostic variants, 152 were unique: 109 heterozygous, 10 compound heterozygous pairs, 18 homozygous, 3 hemizygous, and 2 mosaic variants. One infant had two genetic diagnoses. Most variants were single nucleotide variants or small insertions-deletions (74 missense, 23 frameshift, 26 nonsense, 8 intronic, 2 in-frame indels), 17 were copy number variants (4 intragenic), and 2 repeat expansions.

**Table 1:**
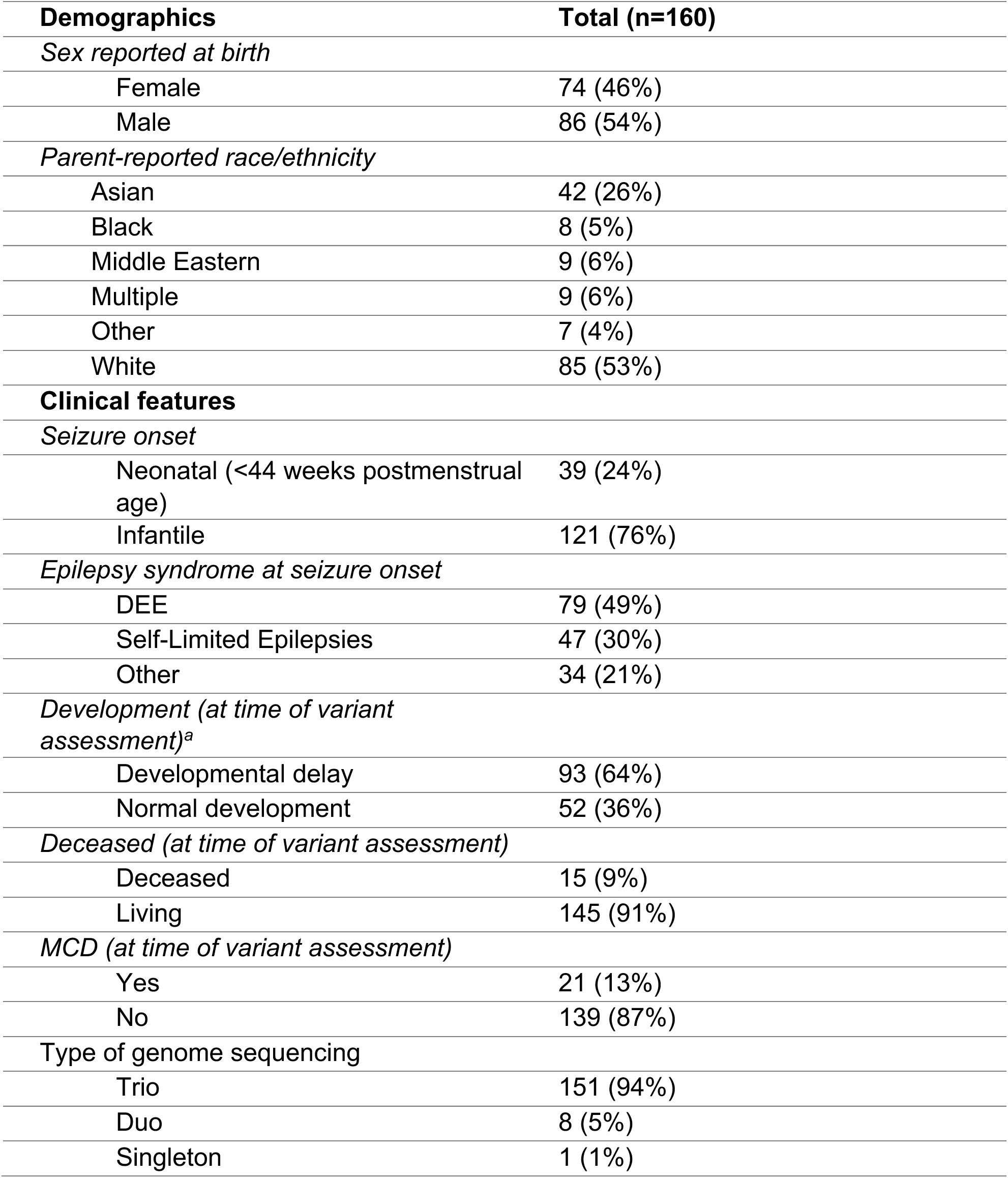
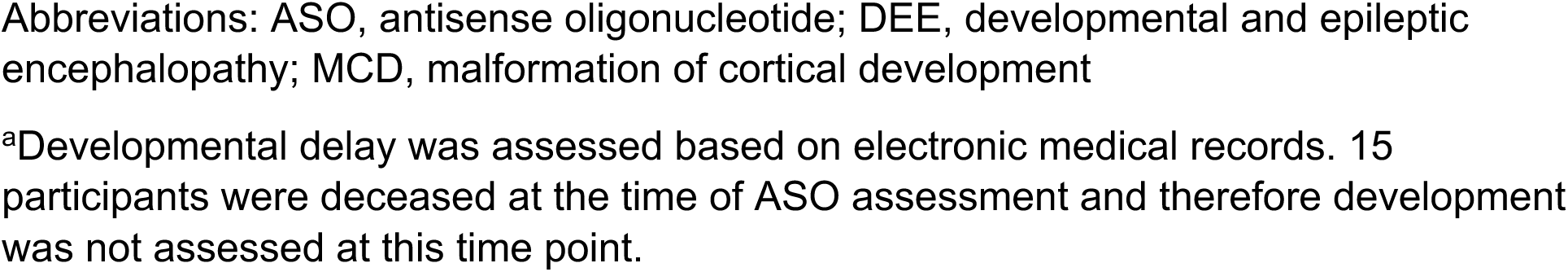
Cohort demographic and clinical characteristics.

| **Demographics** | **Total (n=160)** |
| --- | --- |
| *Sex reported at birth* |  |
| Female | 74 (46%) |
| Male | 86 (54%) |
| *Parent-reported race/ethnicity* |  |
| Asian | 42 (26%) |
| Black | 8 (5%) |
| Middle Eastern | 9 (6%) |
| Multiple | 9 (6%) |
| Other | 7 (4%) |
| White | 85 (53%) |
| **Clinical features** |  |
| *Seizure onset* |  |
| Neonatal (<44 weeks postmenstrual age) | 39 (24%) |
| Infantile | 121 (76%) |
| *Epilepsy syndrome at seizure onset* |  |
| DEE | 79 (49%) |
| Self-Limited Epilepsies | 47 (30%) |
| Other | 34 (21%) |
| *Development (at time of variant assessment)<sup>a</sup>* |  |
| Developmental delay | 93 (64%) |
| Normal development | 52 (36%) |
| *Deceased (at time of variant assessment)* |  |
| Deceased | 15 (9%) |
| Living | 145 (91%) |
| *MCD (at time of variant assessment)* |  |
| Yes | 21 (13%) |
| No | 139 (87%) |
| *Type of genome sequencing* |  |
| Trio | 151 (94%) |
| Duo | 8 (5%) |
| Singleton | 1 (1%) |

Abbreviations: ASO, antisense oligonucleotide; DEE, developmental and epileptic encephalopathy; MCD, malformation of cortical development
<sup>a</sup>Developmental delay was assessed based on electronic medical records. 15 participants were deceased at the time of ASO assessment and therefore development was not assessed at this time point.

N1C guideline assessment classified 15/152 variants (9.9%) as “eligible” for various ASO therapy approaches, 7 (4.6%) “likely eligible”, 12 (7.9%) “unlikely eligible”, 61 (40.1%) “not eligible”, and the remaining 57 (37.5%) “unable to assess”. Splice-switching framework assessment classified one variant as “probably” (this variant was “not eligible” by N1C guidelines) and two “possibly” (one “likely eligible” for splice correction and one “unlikely eligible” by N1C guidelines) amenable to splice correction ASO therapy approaches (**Figure 2A, eTable 1**).

**Figure 2:**
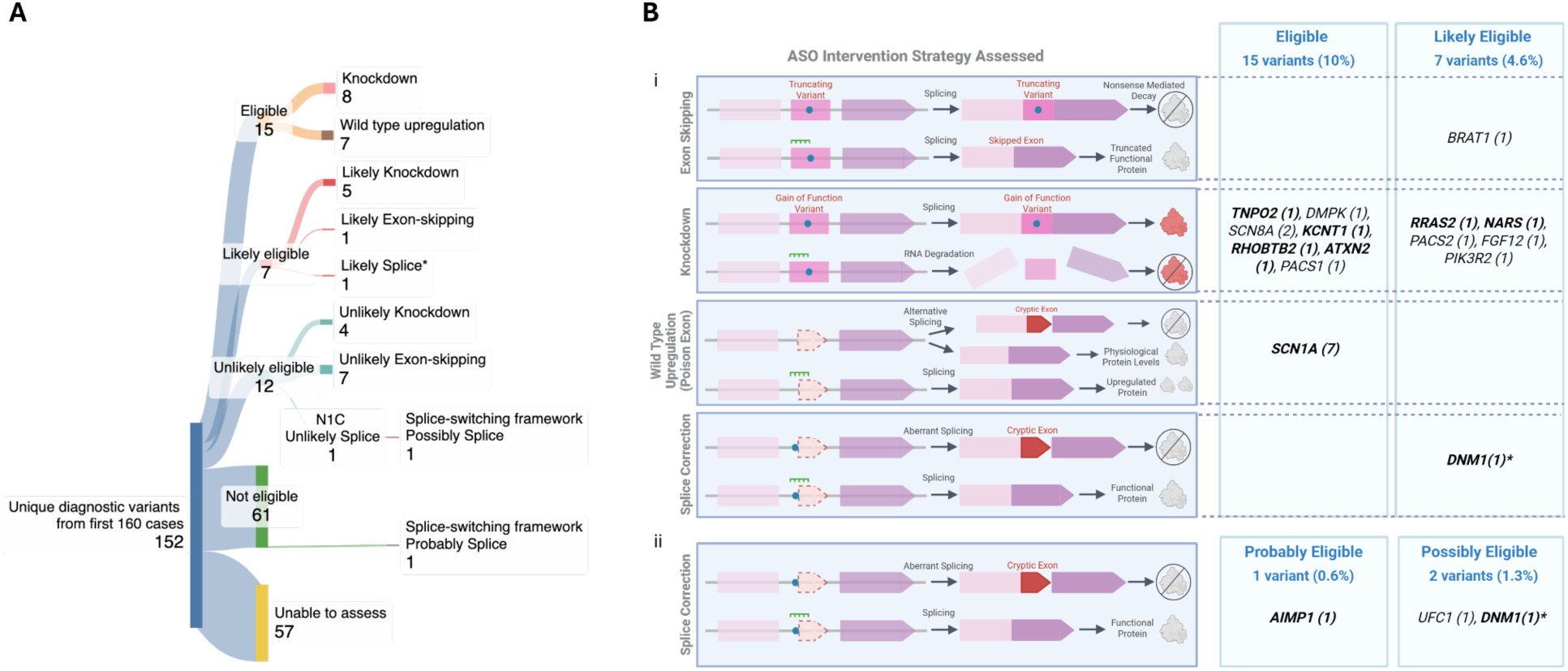
Summary of ASO eligibility assessment results. A) Overview of results for all variants. * indicates variant that was also possibly eligible under the splice-switching framework. B) Summary of results for eligible cases based upon the i) N1C VARIANT guidelines^12^ and ii) splice-switching framework.^11^ The number of variants identified per gene is indicated in brackets. Genes in bold indicate that we would consider ASO intervention based on the patient’s current clinical status. The *KCNT1* variant eligible for knockdown was present in two patients.

Overall, 24 unique variants from 25 infants were in principle eligible for ASO therapy approaches, an eligibility rate of 15.6% (25/160 infants). Of these variants, 13 (54%) were amenable to a knockdown approach, 7 (29%) to wild-type upregulation, 3 (13%) to splice correction, and 1 (4%) to exon-skipping (**Figure 2B**).

Of these 25 infants, 15 (60%) presented with DEEs, 8 (32%) with other syndromes (one evolved to DEE), and 2 (8%) with GEFS+ that evolved to DEEs. Taking into account disease and phenotype considerations, 16/25 infants (64%) could be currently considered for ASO therapy approaches and an additional 5 (20%) could have been considered at seizure onset (3 deceased, 2 with improving trajectory). Of the remaining four infants, two have mild disease severity, one has a brain malformation, and one could be considered in the future if severity worsens.

Of 61 variants classified as “not eligible”, 22 (36%) were autosomal dominant loss-of-function variants in genes possibly eligible for a wild-type upregulation ASO approach, but per N1C guidelines do not have enough data to confidently assess (**eMethods, eTable1**).

Of 57 variants classified as “unable to assess”, largely due to unknown variant pathomechanism, 16 (28%) were in genes targeted by an existing ASO (*KCNT1, SCN1A, SCN2A, SCN8A*). Eight of 17 infants with these variants exhibited phenotypes consistent with the pathomechanism amenable to existing ASO approaches and could in principle be currently considered for ASO therapies given their clinical trajectory.

## Discussion

There is an urgent need to bridge the gap from precision diagnoses to precision therapies for rare genetic diseases. In our cohort of infants with genetic epilepsies, 15.6% (25/160) were in principle eligible for ASO therapy development after applying two published guidelines for variant assessment. Taking into account disease and phenotype considerations, most (21/25 (84%)) could be considered currently or at seizure onset for ASO therapy development, highlighting the potential of such therapies in this population.

Previous studies assessing ASO amenability across broad and sometimes highly selected cohorts have reported eligibility rates from 5%-18%, largely reflecting differences in cohort composition and ASO assessment methodologies.^13, 14^ Our prospectively recruited, rigorously phenotyped infantile epilepsy cohort that underwent first-line GS provides unique insights into the potential of ASO therapies in a disease-specific population. We utilized stringent guidelines that emphasize experimental evidence to ensure the appropriate disease mechanism is targeted. A notable limitation is the 37.5% of variants classified as “unable to assess” by N1C guidelines due to lack of functional evidence validating variant pathomechanism or splicing impact—often the case for rare or novel variants. Validating variant mechanisms is especially crucial to determine eligibility for available ASOs, which eight infants could be eligible for if their variant mechanisms were known. Developing rapid, scalable validation methods for variant pathomechanisms is an important future direction.

Intervention with precision therapies as early as possible in the disease course is key to slowing or preventing disease progression and optimizing outcomes. Rapid GS is emerging as a first-tier diagnostic test for infantile epilepsies. To maximize benefits of early diagnoses, we need rapid variant assessment and mechanism validation to determine eligibility for precision therapy approaches like ASOs, enabling timely identification of patients suitable for an existing ASO or for whom a novel ASO may be developed. This rapid diagnosis-to-therapy framework will require international partnership between clinicians, researchers, laboratories, industry, and regulators to meaningfully and equitably advance precision medicine for infantile epilepsies and rare diseases more broadly.

## Data Access

AMD had full access to all the data in the study and takes responsibility for the integrity of the data and the accuracy of the data analysis.

## Supporting information

eMethods

eTable 1

## Data Availability

Assessed variants are available in eTable 1 and were deposited into public databases (e.g., ClinVar) per the policies of the clinically accredited laboratories who reported the variants.

## Gene-STEPS Study Group

Joanna Cobb, Anna J.S. Griffiths, Edward J. Higgenbotham, Puneet Jain, Nicole S.Y. Liang, Sebastian Lunke, Christian R. Marshall, Catherine Marx, Lyndsey McRae, Jimmy N.H. Nguyen, Wanqing Shao, Beth R. Sheidley, Lacey Smith, Zornitza Stark, Susan M. White.

## Additional Acknowledgements

We thank the patients and their families who participated in the study. We thank the IPCHiP Executive Committee (Nancy C. Andrews, Alan H. Beggs, John Christodoulou, J Helen Cross, Christian R. Marshall, Kathryn N. North, Stephen W. Scherer, Neil J. Sebire, Piotr Sliz) for their support. We also appreciate the ongoing work of the N=1 Collaborative, n-Lorem Foundation, and other groups working in the precision ASO therapy field.

