## Supplementary material for "Assessing Precision Antisense Oligonucleotide Therapy Eligibility for Infantile Genetic Epilepsies": eMethods

### Supplemental File

eMethods 2

eTable 1 4

eReferences 5

### eMethods

#### Study Design and Cohort

The International Precision Child Health Partnership (IPCHiP) is a consortium of four pediatric referral centers: Melbourne Children's Campus (Murdoch Children's Research Institute and The Royal Children's Hospital) in Australia; The Hospital for Sick Children (SickKids) in Canada; University College London Great Ormond Street Institute of Child Health in the United Kingdom; and Boston Children's Hospital in the United States.[1]

Gene-STEPS is a flagship project of IPCHiP. As previously described, the Gene-STEPS study recruits infants with new-onset epilepsy or complex febrile seizures who present to one of the four participating sites.[2] Inclusion criteria include seizure onset <12 months old and enrollment within 6 weeks of site presentation.[2] Exclusion criteria include simple febrile seizures, acute provoked seizures, or known genetic or acquired cause of seizures.[2]

Enrolled infants receive rapid short-read genome sequencing (duo or trio sequencing whenever one or both biological parents are available, respectively) at clinically accredited laboratories. The genome sequencing data is transferred to the study teams for further analysis, with any additional diagnostic variants identified by the study teams confirmed at clinically accredited laboratories.

As previously described,[2] infants were considered genetically diagnosed if (1) pathogenic/likely pathogenic variant(s) were identified in a gene with an appropriate mode of disease inheritance and with a disease association that fit the infant's presentation or (2) variant(s) of uncertain significance (VUS) were identified that were considered clinically diagnostic by expert consensus when additional evidence (e.g., genotype-phenotype correlation, further investigations) supported pathogenicity and no evidence ruled out pathogenicity.

#### Clinical Data

Demographic and clinical data were collected from the electronic medical record (EMR) at enrollment, including sex assigned at birth, parent-reported race/ethnicity, referral setting (neonatal or pediatric intensive care unit, non-intensive care inpatient unit, outpatient clinic), gestational age at birth, epilepsy details (age of seizure onset, seizure types, EEG findings), brain MRI findings, developmental status (developmental delay before seizure onset and/or regression after seizure onset), other neurological and non-neurological features, other genetic testing, and family medical history.

Additional data were collected at the time of ASO assessment, including any change in seizure type or epilepsy syndrome, seizure frequency and presence of drug-resistant epilepsy, anti-seizure medications or other treatments, presence of developmental delay or regression, any other new neurological or non-neurological abnormalities, and if applicable, age at death.

Epilepsy syndromes at onset and at time of assessment were classified based on International League Against Epilepsy definitions.[3]

#### Eligibility Assessment

The N=1 Collaborative (N1C) VARIANT guidelines evaluate and classify variants for amenability to three ASO strategies: splice correction (correction of aberrant splicing causing disease), exon skipping (skipping of canonical exons containing the pathogenic variant), and downregulation of RNA transcripts ("knockdown"). Variants are categorized as "eligible", "likely eligible", "unlikely eligible", "not eligible", or "unable to assess". "Eligible" variants are those for which functional evidence supports the effectiveness of an ASO approach. "Likely eligible" variants meet molecular criteria for ASO development but lack supporting functional evidence. "Unlikely eligible" variants exhibit molecular features that suggest limited ASO feasibility, but no evidence directly contradicts their use. “Not eligible” variants are those for which ASO therapies are not currently feasible due to constraints of ASO technology or evidence demonstrating their ineffectiveness. Variants classified as “unable to assess” have insufficient data to properly assess for ASO amenability or are outside the scope of current guidelines to determine eligibility. Full methodological details are provided in the N1C VARIANT guidelines.[4] Variant classifications are based on the variant and ASO information available at assessment, and can change over time.

The N1C VARIANT guidelines outline considerations for wildtype upregulation strategies but do not include specific criteria for classifying amenability to these strategies. Variants were deemed “possibly eligible” for wildtype upregulation if they were: (i) not eligible for exon skipping, knockdown, or splice correction, (ii) autosomal dominant loss-of-function variants in patients retaining a wildtype allele; and, (iii) impacting a gene annotated as having a potentially targetable poison exon, naturally occurring antisense transcript, or upstream open reading frame, as compiled in the supplemental table of the guideline publications,[4] which aggregates findings from prior studies.[5-8]

For variants classified as “unable to assess”, sufficient evidence of the variant pathomechanism was not available to evaluate ASO amenability. In these cases, where the variant occurred in a gene with a clinically available or pre-clinically validated ASO, patient phenotypes were reviewed to infer the likely pathomechanism based on established genotype-phenotype correlations, and to determine whether the patient phenotype may be suitable for treatment consideration.

In addition to the N1C VARIANT Guidelines, variants were assessed using a previously published splice-switching framework.[9] The splice-switching framework uses a taxonomy to classify disease-causing variants by their predicted amenability as “probably amenable”, “possibly amenable”, or “unlikely amenable”. The framework considers two questions: 1) functional damage and 2) correctability of mis-splicing. *In silico* variant effect predictors such as REVEL, SpliceAI, MaxENT scan, LaBranchoR, and any available functional data are used to assess functional damage.[10-13] Assessment of the correctability of mis-splicing is based on the observation that variants creating novel splice sites are generally more amenable to rescue than those weakening canonical splice sites. Unlike the N1C VARIANT Guidelines, the splice-switching framework incorporates *in silico* predictions into its assessments. Therefore, this framework may classify variants as “probably” or “possibly” eligible using *in silico* predictors despite those variants being classified as “unable to assess” by the N1C VARIANT Guidelines because of a lack of functional evidence.

Patients with variants identified by either or both guidelines as being targetable by an ASO (“eligible” or “likely eligible” according to N1C VARIANT guidelines and/or “probably amenable” or “possibly amenable” according to the splice-switching framework) were further assessed for general disease factors and patient-specific phenotypes. These considerations included seizure/epilepsy details (seizure type, seizure frequency, epilepsy syndrome, response to anti-seizure medications, presence of drug-resistant epilepsy), developmental status, number of emergency room visits and hospitalizations, other neurological and non-neurological comorbidities, and current and predicted future disease trajectory. Each patient underwent multi-site clinician discussion of potential consideration for ASO therapy approach at the time of assessment and/or disease onset using previously described methods.[14, 15]

### eTable 1

Please see the eTable 1 file.
