## Supplementary material for "Assessing Precision Antisense Oligonucleotide Therapy Eligibility for Infantile Genetic Epilepsies": eTable 1

| Participant | Gene/Genomic Region | Transcript | Chromosome cooordinates (GRCh38) | c. notation | p. notation | Variant type | Inheritance | Zygosity | Pathomechanism | N1C VARIANT outcome | Variants that could be eligible for ASO by N1C guidelines if pathomechanism was proven (existing ASO) | Variants that may be suitable for a TANGO approach | Individualized splice switching framework outcome |
|---|---|---|---|---|---|---|---|---|---|---|---|---|---|
| A001 | TSC2 | NM_000548.3 | chr16:2080365C>T | c.3598C>T | p.(Arg1200Trp) | Missense | AD | Heterozygous | LoF | Unlikely eligible for exon skipping | | | Unlikely amenable |
| A002 | PRRT2 | NM_145239.2 | chr16:29813694dup | c.649dupC | p.(Arg217Profs*8) | Frameshift | AD | Heterozygous | LoF | Not eligible | | Possibly for wild type upregulation | Unlikely amenable |
| A003 | 18p11.32p11.21 del | | chr18:11358-15286251del | | | Copy number loss | AD | Heterozygous | N/A | Unable to assess | | | Unlikely amenable |
| A004 | 2q24.1q24.3 dup | | chr2:155579940-166902498dup | | | Copy number gain | AD | Heterozygous | N/A | Unable to assess | | | Unlikely amenable |
| A005 | NSF | NM_006178.3 | chr17:46711081-46711083dup | c.1590_1592dupCAG | p(Ser531dup) | Duplication | AD | Heterozygous | Unknown | Unable to assess | | | Unlikely amenable |
| A006 | RRAS2 | NM_012250.5 | chr11:14358803C>T | c.68G>A | p.(Gly23Asp) | Missense | AD | Heterozygous | GoF | **Likely eligible for knockdown** | | | Unlikely amenable |
| A007 | 16p11.2 del | | chr16:29621833-30188698del | | | Copy number loss | AD | Heterozygous | N/A | Unable to assess | | | Unlikely amenable |
| A008 | SCN1A | NM_001165963.2 | chr2:165992408C>G | c.4867G>C | p.(Glu1623Gln) | Missense | AD | Heterozygous | Unknown (likely LoF by phenotype) | Unable to assess | **Would be eligible for WT upregulation** | | Unlikely amenable |
| A009 | TUBA1A | NM_001270399.1 | chr12:49186666-49186669delinsAGCATTCAGAGAT | c.168_171delinsAGCATTCAGAGAT | p.(Gly57delinsAlaPheArgAsp) | Insertion/deletion | AD | Heterozygous | Unknown | Unable to assess | | | Unlikely amenable |
| A010 | PRRT2 | NM_145239.2 | chr16:29813694dup | c.649dupC | p.(Arg217Profs*8) | Frameshift | AD | Heterozygous | LoF | Not eligible | | Possibly for wild type upregulation | Unlikely amenable |
| A011 | SCN8A | NM_001330260.1 | chr12:51786566G>A | c.3967G>A | p.(Ala1323Thr) | Missense | AD | Heterozygous | Unknown (likely GoF by phenotype) | Unable to assess | **Would be eligible for knockdown** | | Unlikely amenable |
| A012 | PRRT2 | NM_145239.2 | chr16:29813694dup | c.649dupC | p.(Arg217Profs*8) | Frameshift | AD | Heterozygous | LoF | Not eligible | | Possibly for wild type upregulation | Unlikely amenable |
| A013 | SCN2A | NM_001040142.1 | chr2:165373331G>C | c.3956G>C | p.(Arg1319Pro) | Missense | AD | Heterozygous | Unknown (likely GoF by phenotype) | Unable to assess | **Would be eligible for knockdown** | | Unlikely amenable |
| A014 | SCN1B | NM_001037.4 | chr19:35033654C>G | c.363C>G | p.(Cys121Trp) | Missense | AR | Compound heterozygous | LoF | Not eligible | | | Unlikely amenable |
| A014 | SCN1B | NM_001037.4 | chr19:35032665C>T | c.178C>T | p.(Arg60Cys) | Missense | AR | Compound heterozygous | LoF | Not eligible | | | Unlikely amenable |
| A015 | CSNK2B | NM_001320.6 | chr6:31669359C>G | c.408C>G | p.(Tyr136*) | Nonsense | AD | Heterozygous | LoF | Not eligible | | | Unlikely amenable |
| A016 | TCF4 | NM_001083962.1 | chr18:55228987C>T | c.1739G>A | p.(Arg580Gln) | Missense | AD | Heterozygous | LoF | Not eligible | | Possibly for wild type upregulation | Unlikely amenable |
| A017 | SCN8A | NM_001330260.1 | chr12:51786552A>G | c.3953A>G | p.(Asn1318Ser) | Missense | AD | Heterozygous | Unknown (likely GoF by phenotype) | Unable to assess | **Would be eligible for knockdown** | | Unlikely amenable |
| A018 | ADSL | NM_000026.3 | chr22:40365003C>T | c.1315C>T | p.(Gln439*) | Nonsense | AR | Heterozygous | LoF | Not eligible | | | Unlikely amenable |
| A018 | ADSL | NM_000026.3 | chr22:40358949C>G | c.568C>G | p.(Arg190Gly) | Missense | AR | Heterozygous | LoF | Not eligible | | | Unlikely amenable |
| A019 | NARS | NM_004539.3 | chr18:57601699G>A | c.1600C>T | p.(Arg534*) | Nonsense | AD | Heterozygous | Dominant negative | **Likely eligible for knockdown** | | | Unlikely amenable |
| A020 | SETBP1 | NM_015559.3 | chr18:44951949G>A | c.2609G>A | p.(Gly870Asp) | Missense | AD | Heterozygous | GoF | Unlikely eligible for knockdown | | | Unlikely amenable |
| A021 | KCNT1 | NM_020822.2 | chr9:135778744A>T | c.2651A>T | p.(Asp884Val) | Missense | AD | Heterozygous | Unknown (likely GoF by phenotype) | Unable to assess | **Would be eligible for knockdown** | | Unlikely amenable |
| A022 | HSD17B4 | NM_000414.3 | chr5:119493838G>A | c.760G>A | p.(Gly254Arg) | Missense | AR | Homozygous | LoF | Not eligible | | | Unlikely amenable |
| A023 | STXBP1 | NM_001032221.3 | chr9:127663344G>A | c.569G>A | p.(Arg190Gln) | Missense | AD | Heterozygous | Unknown (likely LoF by phenotype) | Unable to assess | | | Unlikely amenable |
| A024 | PRRT2 | NM_145239.2 | chr16:29813694dup | c.649dupC | p.(Arg217Profs*8) | Frameshift | AD | Heterozygous | LoF | Not eligible | | Possibly for wild type upregulation | Unlikely amenable |
| A025 | MOCS1 | NM_001358530.1 | chr6:39912274C>T | c:971G>A | p.(Gly324Glu) | Missense | AR | Homozygous | LoF | Not eligible | | | Unlikely amenable |
| A026 | ACO2 | NM_001098.2 | chr22:41528607-41528608del | c.2338_2339del | p.(Gln780Valfs*63) | Frameshift | AR | Homozygous | LoF | Not eligible | | | Unlikely amenable |
| A027 | CYFIP2 | NM_001037333.2 | chr5:157294834C>T | c.259C>T | p.(Arg87Cys) | Missense | AD | Heterozygous | GoF | Not eligible | | | Unlikely amenable |
| A028 | KCNT1 | NM_020822.2 | chr9:135765706G>A | c.1283G>A | p.(Arg428Gln) | Missense | AD | Heterozygous | GoF | **Eligible for knockdown** | | | Unlikely amenable |
| A029 | KCNQ2 | NM_172107.3 | chr20:63415086C>T | c.1342C>T | p.(Arg488*) | Nonsense | AD | Heterozygous | LoF | Not eligible | | Possibly for wild type upregulation | Unlikely amenable |
| A030 | 16p11.2 dup | | chr16: 29616028_30188392dup | | | Copy number gain | AD | Heterozygous | N/A | Unable to assess | | | Unlikely amenable |
| A031 | PACS2 | NM_001100913.2 | chr14:105368112G>A | c.625G>A | p.(Glu209Lys) | Missense | AD | Heterozygous | GoF | **Likely eligible for knockdown** | | | Unlikely amenable |
| A032 | UFSP2 | NM_018359.3 | chr4: 185415857A>T | c.344T>A | p.(Val115Glu) | Missense | AR | Homozygous | LoF | Not eligible | | | Unlikely amenable |
| A033 | PRRT2 | NM_145239.2 | chr16:29813703C>T | c.649C>T | p.(Arg217*) | Nonsense | AD | Heterozygous | LoF | Not eligible | | Possibly for wild type upregulation | Unlikely amenable |
| A034 | PRRT2 | NM_145239.2 | chr16:29813695del | c.649del | p.(Arg217Glufs*12) | Frameshift | AD | Heterozygous | LoF | Not eligible | | Possibly for wild type upregulation | Unlikely amenable |
| A035 | CSNK2B | NM_001320.6 | chr6:31669095A>G | c.292-2A>G | | Intronic SNV | AR | Homozygous | Unknown | Unable to assess | | | Unlikely amenable |
| A036 | SCN1A | NM_001165963.2 | chr2:165996047G>T | c.4547C>A | p.(Ser1516*) | Nonsense | AD | Heterozygous | LoF | **Eligible for WT upregulation** | | | Unlikely amenable |
| A037 | PRRT2 | NM_145239.2 | chr16:29813694dup | c.649dupC | p.(Arg217Profs*8) | Frameshift | AD | Heterozygous | LoF | Not eligible | | Possibly for wild type upregulation | Unlikely amenable |
| A038 | PRRT2 | NM_145239.2 | chr16:29813694dup | c.649dupC | p.(Arg217Profs*8) | Frameshift | AD | Heterozygous | LoF | Not eligible | | Possibly for wild type upregulation | Unlikely amenable |
| A039 | KCNH5 | NM_139318.4 | chr14:62950522G>A | c.980G>A | p.(Arg327His) | Missense | AD | Heterozygous | GoF | Unlikely eligible for knockdown | | | Unlikely amenable |
| A040 | SCN1A | NM_001165963.2 | chr2:165991672_165991673insGGAT | c.5602_5603insATCC | p.(Leu1868Tyrfs*78) | Frameshift | AD | Heterozygous | LoF | **Eligible for WT upregulation** | | | Unlikely amenable |
| A041 | DEPDC5 | NM_001242896.1 | chr22:31846873del | c.3061del | p.(Ile1021Phefs*58) | Frameshift | AD | Heterozygous | LoF | Not eligible | | Possibly for wild type upregulation | Unlikely amenable |
| A042 | DYNC1H1 | NM_001376.4 | chr14:102012445G>A | c.6989G>A | p.(Gly2330Glu) | Missense | AD | Heterozygous | Unknown | Unable to assess | | | Unlikely amenable |
| A043 | TNPO2 | NM_001136196.1 | chr19:12715505C>T | c.466G>A | p.(Asp156Asn) | Missense | AD | Heterozygous | GoF | **Eligible for knockdown** | | | Unlikely amenable |
| A044 | PPP3CA | NM_000944.4 | chr4:101032353-101032354del | c.1255_1256del | p.(Ser419Cysfs*31) | Frameshift | AD | Heterozygous | LoF | Not eligible | | Possibly for wild type upregulation | Unlikely amenable |
| A045 | PTEN | NM_000314.4 | chr10:87864518C>T | c.49C>T | p.(Gln17*) | Nonsense | AD | Heterozygous | LoF | Not eligible | | | Unlikely amenable |
| A046 | 2q24.2q24.3 del | | chr2:160825591-166146619del | | | Copy number loss | AD | Heterozygous | N/A | Unable to assess | | | Unlikely amenable |
| A047 | DMPK | NM_004409.5 | | c.*224_226CTG[>200]; confirmed >2000 on subsequent Southern blot | | Repeat expansion | AD | Heterozygous | N/A | **Eligible for knockdown** | | | Unlikely amenable |
| A048 | SCN8A | NM_014191.3 | chr12:51780755G>A | c.3926G>A | p.(Arg1309Gln) | Missense | AD | Heterozygous | Unknown (likely GoF by phenotype) | Unable to assess | **Would be eligible for knockdown** | | Unlikely amenable |
| A049 | SETD5 | NM_001080517.1 | chr3:9447870T>G | c.1967T>G | p.(Leu656*) | Nonsense | AD | Heterozygous | LoF | Unlikely eligible for exon-skipping | | | Unlikely amenable |
| A050 | ZC4H2 | NM_018684.3 | chrX:64919158del | c.450del | p.(Ile151Serfs*36) | Frameshift | X-linked D | Heterozygous | LoF | Not eligible | | | Unlikely amenable |
| A051 | SCN2A | NM_001040143.1 | chr2:165309221T>C | c.662T>C | p.(Val221Ala) | Missense | AD | Heterozygous | Unknown (likely GoF by phenotype) | Unable to assess | **Would be eligible for knockdown** | | Unlikely amenable |
| A052 | MOGS | NM_006302.2 | chr2:74462328C>G | c.1461G>C | p.(Glu487Asp) | Missense | AR | Compound heterozygous | LoF | Unable to assess | | | Unlikely amenable |
| A052 | MOGS | NM_006302.2 | chr2:74461319C>T | c.2470G>A | p.(Gly824Ser) | Missense | AR | Compound heterozygous | LoF | Unable to assess | | | Unlikely amenable |
| A053 | MOCS2 | NM_004531.5 | chr5:53102097C>T | c.226G>A (reported as NM_176806.2: c.*146 G>A) | p.(Gly76Arg) | Missense | AR | Homozygous | LoF | Unable to assess | | | Unlikely amenable |
| A054 | CSNK2B | NM_001320.6 | chr6:31669508G>A | c.557G>A | p.(Arg186Lys) | Missense | AD | Heterozygous | LoF | Unable to assess | | | Unlikely amenable |
| A055 | SCN2A | NM_021007.2 | chr2:165389093T>A | c.5287T>A | p.(Ser1763Thr) | Missense | AD | Heterozygous | Unknown (likely GoF by phenotype) | Unable to assess | **Would be eligible for knockdown** | | Unlikely amenable |

| | | | | | | | | | | | | | |
|---|---|---|---|---|---|---|---|---|---|---|---|---|---|
| A056 | UGDH | NM_003359.3 | chr4:39514154G>A | c.193C>T | p.(Arg65Ter) | Missense | AR | Compound heterozygous | LoF | Unlikely eligible for exon-skipping | | | Unlikely amenable |
| A056 | UGDH | NM_003359.3 | chr4:39509862T>C | c.709A>G | p.(Ile237Val) | Missense | AR | Compound heterozygous | LoF | Unable to assess | | | Unlikely amenable |
| A057 | PRRT2 | NM_145239.2 | chr16:29813703dup | c.649dup | p.(Arg217Profs*8) | Frameshift | AD | Heterozygous | LoF | Not eligible | | Possibly for wild type upregulation | Unlikely amenable |
| A058 | 1p36.33p36.32 del | | chr1:817963-3077912del | | | Copy number loss | AD | Heterozygous | N/A | Unable to assess | | | Unlikely amenable |
| A059 | WWOX | | chr16:78424000-78558906del | | | intragenic deletion | AR | Compound heterozygous | N/A | Not eligible | | | Unlikely amenable |
| A059 | WWOX | | chr16:79114704-79266751del | | | intragenic deletion | AR | Compound heterozygous | N/A | Not eligible | | | Unlikely amenable |
| A060 | PRRT2 | NM_145239.2 | chr16:29813703dup | c.649dup | p.(Arg217Profs*8) | Frameshift | AD | Heterozygous | LoF | Not eligible | | Possibly for wild type upregulation | Unlikely amenable |
| A061 | PRRT2 | NM_145239.2 | chr16:29813267-29813268del | c.215_216del | p.(Thr72Argfs*61) | Frameshift | AD | Heterozygous | LoF | Not eligible | | Possibly for wild type upregulation | Unlikely amenable |
| A062 | SCN8A | NM_014191.3 | chr12:51807116A>G | c.5630A>G | p.(Asn1877Ser) | Missense | AD | Heterozygous | GoF | **Eligible for knockdown** | | | Unlikely amenable |
| A063 | KCNQ2 | NM_014191.3 | chr20:63433815_63433818dup | c.1109_1112dup | p.(Met371Ilefs*31) | Frameshift | AD | Heterozygous | LoF | Not eligible | | Possibly for wild type upregulation | Unlikely amenable |
| A064 | SCN1A | NM_014191.3 | chr2:166046969C>A | c.1178G>T | p.(Arg393Leu) | Missense | AD | Heterozygous | Unknown (likely LoF by phenotype) | Unable to assess | **Would be eligible for WT upregulation** | | Unlikely amenable |
| A065 | 15q11.2q13.2 dup | | hg19: chr15:22832965_30371538 gain, couldn't convert with liftover | | | Copy number gain | AD | Heterozygous | N/A | Unable to assess | | | Unlikely amenable |
| A066 | NPRL3 | NM_001077350.2 | chr16:119154G>C | c.290C>G | p.(Pro97Arg) | Missense | AD | Heterozygous | Unknown | Unable to assess | | | Unlikely amenable |
| A067 | L1CAM | NM_000425.3 | chrX:153866792C>G | c.2288G>C | p.(Trp763Ser) | Missense | X-linked R | Heterozygous | Unknown | Unable to assess | | | Unlikely amenable |
| A068 | KANSL1 | NM_001193466.1 | chr17:46170911dup | c.1233dup | p.(Asp412Ter) | Missense | AD | Heterozygous | LoF | Not eligible | | Possibly for wild type upregulation | Unlikely amenable |
| A069 | NEXMIF | NM_001008537.1 | chrX:74743593G>A | c.964 C>T | p.(Arg322Ter) | Missense | X-linked D | Hemizygous | LoF | Not eligible | | | Unlikely amenable |
| A070 | PRRT2 | NM_145239.2 | chr16:29813926C>T | c.872C>T | p.(Ala291Val) | Missense | AD | Heterozygous | Unknown | Unable to assess | | | Unlikely amenable |
| A071 | KMT2D | NM_003482.3 | chr12:49038452dup | c.8903dup | p.(Ser2969Valfs*4) | Frameshift | AD | Heterozygous | Unknown | Unable to assess | | | Unlikely amenable |
| A072 | GRIA1 | NM_001114183.1 | chr5:153677161G>C | c.1029G>C | p.(Gln343His) | Missense | AD | Heterozygous | Unknown | Unable to assess | | | Unlikely amenable |
| A072 | PRRT2 | NM_145239.2 | chr16:29813703dup | c.649dup | p.(Arg217Profs*8) | Frameshift | AD | Heterozygous | LoF | Not eligible | | Possibly for wild type upregulation | Unlikely amenable |
| A073 | 20q13.33 del | | chr20:63443194-63693395del | | | Copy number loss | AD | Heterozygous | N/A | Unable to assess | | | Unlikely amenable |
| A074 | SCN2A | NM_021007.2 | chr2:165389093T>A | c.5287T>A | p.(Ser1763Thr) | Missense | AD | Heterozygous | Unknown (likely GoF by phenotype) | Unable to assess | **Would be eligible for knockdown** | | Unlikely amenable |
| A075 | PRUNE1 | NM_021222.1 | chr1:151025367-151026816 | c.521-148_680-417del | | Intragenic deletion | AR | Homozygous | N/A | Not eligible | | | Unlikely amenable |
| A076 | ST3GAL5 | NM_003896.3 | chr2:85840338C>T | c.1063G>A | p.(Glu355Lys) | Missense | AR | Homozygous | LoF | Not eligible | | | Unlikely amenable |
| A077 | TBCK | NM_001163435.2 | chr4:106262103G>A | c.376C>T | p.(R126*) | Nonsense | AR | Homozygous | LoF | Not eligible | | | Unlikely amenable |
| A078 | POU3F3 | NM_006236.1 | chr2:104856167-104856168del | c.658_659del | p.(L220Afs*?) | Frameshift | AD | Heterozygous | LoF | Not eligible | | Possibly for wild type upregulation | Unlikely amenable |
| A079 | DBT | NM_001918.2 | chr1:100216085C>A | c.670 G>T | p.(Glu224Ter) | Missense | AR | Compound heterozygous | LoF | Not eligible | | | Unlikely amenable |
| A079 | DBT | NM_001918.2 | chr1:100240885C>T | c.52-1 G>A | p.? | Intronic SNV | AR | Compound heterozygous | LoF | Not eligible | | | Unlikely amenable |
| A080 | TCF4 | | chr18:55092493-55657706del | | | Copy number loss | AD | Heterozygous | LoF | Not eligible | | Possibly for wild type upregulation | Unlikely amenable |
| A081 | KCNQ2 | NM_172107.4 | chr20:g.63407313delG | c.1955del | p.(Pro652Argfs*278) | Frameshift | AD | Heterozygous | Unknown | Unable to assess | | | Unlikely amenable |
| A082 | PRRT2 | NM_145239.3 | chr16:g.29813703C>T | c.649C>T | p.(Arg217*) | Nonsense | AD | Heterozygous | LoF | Not eligible | | Possibly for wild type upregulation | Unlikely amenable |
| A083 | DEAF1 | NM_021008.4 | chr11:g.686953G>T | c.709C>A | p.(Pro237Thr) | Missense | AD | Heterozygous | Unknown | Unable to assess | | | Unlikely amenable |
| A084 | SCN8A | NM_014191.4 | chr12:g.51790419A>G | c.4441A>G | p.(Met1481Val) | Missense | AD | Heterozygous | Unknown (likely GoF by phenotype) | Unable to assess | **Would be eligible for knockdown** | | Unlikely amenable |
| A085 | 16p11.2 del | | chr16:29652060-30199344del | | | Copy number loss | AD | Heterozygous | N/A | Unable to assess | | | Unlikely amenable |
| A086 | SCN2A | NM_021007.3 | chr2:g.165307876A>G | c.415A>G | p.(Ile139Val) | Missense | AD | Heterozygous | Unknown (likely GoF by phenotype) | Unable to assess | **Would be eligible for knockdown** | | Unlikely amenable |
| A087 | KCNQ2 | NM_172107.4 | chr20:g.63439684C>T | c.841G>A | p.(Gly281Arg) | Missense | AD | Heterozygous | DN | **Unlikely eligible for knockdown** | | | Unlikely amenable |
| A088 | SLC6A5 | NM_004211.5 | chr11:g.20601421_20601422delAG | c.296_297del | p.(Glu99Glyfs*22) | Frameshift | AR | Homozygous | LoF | **Unlikely eligible for exon skipping** | | | Unlikely amenable |
| A089 | KCNQ3 | NM_004519.4 | chr8:g.132174295G>A | c.988C>T | p.(Arg330Cys) | Missense | AD | Heterozygous | LoF | Not eligible | | Possibly for wild type upregulation | Unlikely amenable |
| A090 | RHOBTB2 | NM_001160036.2 | chr8:g.23007627G>A | c.1448G>A | p.(Arg483His) | Missense | AD | Heterozygous | GoF | **Eligible for knockdown** | | | Unlikely amenable |
| A091 | PRRT2 | NM_145239.3 | chr16:g.29813703dupC | c.649dup | p.(Arg217Profs*8) | Frameshift | AD | Heterozygous | LoF | Not eligible | | Possibly for wild type upregulation | Unlikely amenable |
| A092 | KCNT1 | NM_020822.3 | chr9:g.135768922C>A | c.1495C>A | p.(His499Asn) | Missense | AD | Heterozygous | Unknown (likely GoF by phenotype) | Unable to assess | **Would be eligible for knockdown** | | Unlikely amenable |
| A093 | SCN1A | NM_001165963.3 | chr2:g.166038129G>A | c.2593C>T | p.(Arg865*) | Nonsense | AD | Heterozygous | LoF | **Eligible for WT upregulation** | | | Unlikely amenable |
| A094 | HSD17B4 | NM_000414.4 | chr5:g.119489282A>T | c.713A>T | p.(Glu238Val) | Missense | AR | Homozygous | LoF | Not eligible | | | Unlikely amenable |
| A095 | PRRT2 | NM_145239.3 | chr16:g.29813703dupC | c.649dup | p.(Arg217Profs*8) | Frameshift | AD | Heterozygous | LoF | Not eligible | | Possibly for wild type upregulation | Unlikely amenable |
| A096 | FGF12 | NM_021032.4 | chr3:g.192335434C>T | c.341G>A | p.(Arg114His) | Missense | AD | Heterozygous | GoF | **Likely Eligible for knockdown** | | | Unlikely amenable |
| A097 | CDKL5 | | chrX:18474185-18579246dup | | | Intragenic duplication | XL | Hemizygous | N/A | Unable to assess | | | Unlikely amenable |
| A098 | ATXN2 | | | CAG repeat expansion (exon 1, greater than 100 repeats) | | Repeat expansion | AD | Heterozygous | GoF | **Eligible for knockdown** | | | Unlikely amenable |
| A099 | DEPDC5 | NM_001242896.3 | chr22:g.31879638G>T | c.3919G>T | p.(Glu1307*) | Nonsense | AD | Heterozygous | LoF | Unlikely eligible for exon skipping | | Possibly for wild type upregulation | Unlikely amenable |
| A100 | KCNQ2 | NM_172107.4 | chr20:g.63433884G>A | c.1043C>T | p.(Ala348Val) | Missense | AD | Heterozygous | Unknown | Unable to assess | | | Unlikely amenable |
| A101 | KCNQ2 | NM_172107.4 | chr20:g.63413556G>A | c.1657C>T | p.(Arg553Trp) | Missense | AD | Heterozygous | Unknown | Unable to assess | | | Unlikely amenable |
| A102 | KCNT1 | NM_020822.3 | chr9:g.135765706G>A | c.1283G>A | p.(Arg428Gln) | Missense | AD | Heterozygous | GoF | **Eligible for knockdown** | | | Unlikely amenable |
| A103 | PRRT2 | NM_145239.3 | chr16:g.29813703dupC | c.649dup | p.(Arg217Profs*8) | Frameshift | AD | Heterozygous | LoF | Not eligible | | Possibly for wild type upregulation | Unlikely amenable |
| A104 | SCN1A | NM_001165963.4 | chr2:g.165996101A>G | c.4493T>C | p.(Ile1498Thr) | Missense | AD | Heterozygous | GoF | Unlikely eligible for knockdown | | | Unlikely amenable |
| A105 | 17q12 del | | chr17:g.36455272-37889503del | | | Copy number loss | AD | Heterozygous | N/A | Unable to assess | | | Unlikely amenable |
| A106 | ITPA | NM_033453.4 | chr20:g.3213331delA | c.137del | p.(Gln46Argfs*43) | Deletion | AR | Homozygous | LoF | Not eligible | | | Unlikely amenable |
| A107 | 16p11.2 del | | chr16:29600730-30188250del | | | Copy number loss | AD | Heterozygous | N/A | Unable to assess | | | Unlikely amenable |
| A108 | Unbalanced Translocation of 23 OMIM genes | | | | | Unbalanced translocation | | | N/A | Unable to assess | | | Unlikely amenable |
| A109 | PRRT2 | NM_145239.3 | chr16:g.29813703dupC | c.649dup | p.(Arg217Profs*8) | Frameshift | AD | Heterozygous | LoF | Not eligible | | Possibly for wild type upregulation | Unlikely amenable |
| A110 | AP3B2 | NM_004644.5 | chr15:g.82664881G>T | c.2034C>A | p.(Tyr678*) | Nonsense | AR | Homozygous | LoF | Not eligible | | | Unlikely amenable |

| | | | | | | | | | | | | | |
|---|---|---|---|---|---|---|---|---|---|---|---|---|---|
| A111 | PRRT2 | NM_145239.3 | chr16:g.29813565_29813566 delCT | c.511_512del | p.(Leu171Valfs*2) | Frameshift | AD | Heterozygous | LoF | Not eligible | | Possibly for wild type upregulation | Unlikely amenable |
| A112 | SCN1A | NM_001165963.3 | chr2:g.166012271_16601227 2dupAT | c.3724_3725dup | p.(Asp1243Leufs*28) | Frameshift | AD | Heterozygous | LoF | **Eligible for WT upregulation** | | | Unlikely amenable |
| A113 | CDKL5 | NM_001323289.2 | chrX:g.18628659C>T | c.2785C>T | p.(Gln929*) | Nonsense | XL | Heterozygous | LoF | Not eligible | | | Unlikely amenable |
| A114 | NSD1 | NM_022455.5 | chr5:g.177210519C>G | c.2120C>G | p.(Ser707*) | Nonsense | AD | Heterozygous | LoF | Not eligible | | Possibly for wild type upregulation | Unlikely amenable |
| A115 | KCNQ2 | NM_172107.4 | chr20:g.63442497C>T | c.725G>A | p.(Cys242Tyr) | Missense | AD | Heterozygous | Unknown | Unable to assess | | | Unlikely amenable |
| A116 | RNF213 | NM_001256071.3 | chr17:g.80369794T>C | c.12352T>C | p.(Ser4118Pro) | Missense | AD | Heterozygous | Unknown | Unable to assess | | | Unlikely amenable |
| A117 | PRRT2 | NM_145239.3 | chr16:g.29813675dupA | c.621dup | p.(Ser208Ilefs*17) | Frameshift | AD | Heterozygous | LoF | Not eligible | | Possibly for wild type upregulation | Unlikely amenable |
| A118 | PHF21A | NM_001101802.3 | chr11:g.45934055delA | c.1956del | p.(Ala653Profs*103) | Frameshift | AD | Heterozygous | Unknown | Unable to assess | | | Unlikely amenable |
| A119 | UGP2 | NM_006759.4 | chr2:g.63856320A>G | c.34A>G | p.(Met12Val) | Missense | AR | Homozygous | LoF | Not eligible | | | Unlikely amenable |
| A120 | UFC1 | NM_016406.4 | chr1:161157248T>C | c.192-6C>T | p.? | Intronic SNV | AR | Homozygous | LoF | Unlikely eligible for splice correction | | | **Possibly amenable** |
| A121 | STXBP1 | NM_003165.3 | chr9:g.127660043T>C | c.260T>C | p.(Leu87Pro) | Missense | AD | Heterozygous | Unknown | Unable to assess | | | Unlikely amenable |
| A122 | BRAT1 | NM_001350626.1 | chr7:g.2539548C>T | c.1593G>A | p.(Trp531*) | Nonsense | AR | Compound heterozygous | LoF | **Likely eligible for exon skipping** | | | Unlikely amenable |
| A122 | BRAT1 | NM_001350626.1 | chr7:g.2545044G>GT | c.294dup | p.(Leu99Thrfs*92) | Frameshift | AR | Compound heterozygous | LoF | Not eligible | | | Unlikely amenable |
| A123 | GABRB3 | NM_000814.5 | chr15:g.26772404T>C | c.238A>G | p.(Met80Val) | Missense | AD | Heterozygous | GoF | Not eligible | | | Unlikely amenable |
| A124 | KCNQ2 | NM_172107.2 | chr20:g.63438650C>T | c.998G>A | p.(Arg333Gln) | Missense | AD | Heterozygous | GoF | Unlikely eligible for exon skipping and knockdown | | | Unlikely amenable |
| A125 | POLR3B | NM_018082.5 | chr12:g.106433856G>A | c.1765G>A | p.(Gly589Arg) | Missense | AD | Heterozygous | DN | Not eligible | | | Unlikely amenable |
| A126 | ATP6V1A | NM_001690.3 | chr3:g.113788757G>A | c.761G>A | p.(Cys254Tyr) | Missense | AD | Heterozygous | Unknown | Unable to assess | | | Unlikely amenable |
| A127 | CDKL5 | NM_001323289.2 | chrX:g.18604572C>T | c.1648C>T | p.(Arg550*) | Nonsense | XLD | Heterozygous | LoF | Not eligible | | | Unlikely amenable |
| A128 | STXBP1 | NM_003165.4 | chr9:g.127666205C>T | c.703C>T | p.(Arg235*) | Nonsense | AD | Heterozygous | LoF | Not eligible | | Possibly for wild type upregulation | Unlikely amenable |
| A129 | SCN8A | NM_014191.3 | chr12:g.51807101G>A | c.5615G>A | p.(Arg1872Gln) | Missense | AD | Heterozygous | GoF | **Eligible for knockdown** | | | Unlikely amenable |
| A130 | PIGN | NM_176787.5 | chr18:g.62114560C>T | c.1251+1G>A | p.? | Intronic SNV | AR | Compound heterozygous | LoF | Not eligible | | | Unlikely amenable |
| A130 | PIGN | NM_176787.5 | chr18:g.62146972C>G | c.804G>C | p.(Trp268Cys) | Missense | AR | Compound heterozygous | LoF | Not eligible | | | Unlikely amenable |
| A131 | KCNQ2 | NM_172107.3 | chr20:g.63413534C>T | c.1679G>A | p.(Arg560Gln) | Missense | AD | Heterozygous | Unknown | Unable to assess | | | Unlikely amenable |
| A132 | TCF4 | NM_001083962.2 | chr18:g.55229077C>T | c.1650-1G>A | p.? | Intronic SNV | AD | Heterozygous | Unknown | Unable to assess | | | Unlikely amenable |
| A133 | SCN1A | NM_001165963.4 | chr2:g.166041343G>A | c.2303C>T | p.(Pro768Leu) | Missense | AD | Heterozygous | Unknown (likely LoF by phenotype) | Unable to assess | **Would be eligible for WT upregulation** | | Unlikely amenable |
| A134 | SCN1A | NM_001165963.4 | chr2:g.166045135C>A | c.1570G>T | p.(Glu524*) | Nonsense | AD | Heterozygous | LoF | **Eligible for WT upregulation** | | | Unlikely amenable |
| A135 | ATP1A2 | NM_000702.4 | chr1:g.160139735C>T | c.2936C>T | p.(Pro979Leu) | Missense | AD | Heterozygous | LoF | Unlikely eligible for exon skipping | | | Unlikely amenable |
| A136 | SCN1A | NM_001165963.4 | chr2:g.166009736G>A | c.3985C>T | p.(Arg1329*) | Nonsense | AD | Heterozygous | LoF | **Eligible for WT upregulation** | | | Unlikely amenable |
| A137 | PIK3R2 | NM_005027.3 | chr19:g.18162974G>A | c.1117G>A | p.(Gly373Arg) | Missense | AD | Mosaic | GoF | **Likely eligible for knockdown** | | | Unlikely amenable |
| A138 | DNM1 | NM_001288739.2 | chr9:g.128225994G>A | c.1197-41G>A | p.? | Intronic SNV | AD | Heterozygous | | **Likely eligible for splice correction** | | | **Possibly amenable** |
| A139 | ALG11 | NM_001004127.3 | chr13:g.52024767G>A | c.1037G>A | p.(Arg346His) | Missense | AR | Heterozygous | LoF | Not eligible | | | Unlikely amenable |
| A139 | ALG11 | NM_001004127.3 | chr13:g.52024285T>A | c.555T>A | p.(Phe185Leu) | Missense | AR | Heterozygous | LoF | Not eligible | | | Unlikely amenable |
| A140 | PRRT2 | NM_145239.3 | chr16:g.29813694G>GC | c.649dup | p.(Arg217Profs*8) | Frameshift | AD | Heterozygous | LoF | Not eligible | | Possibly for wild type upregulation | Unlikely amenable |
| A141 | STXBP1 | NM_001032221.6 | chr9:g.127651624AG>A | c.60del | p.(Lys21Argfs*16) | Frameshift | AD | Heterozygous | LoF | Not eligible | | Possibly for wild type upregulation | Unlikely amenable |
| A142 | AIMP1 | NM_001142416.2 | chr4:g.106347598A>G | c.845A>G | p.(Asp282Gly) | Missense | AR | Homozygous | LoF | Not eligible | | | Unlikely amenable |
| A143 | KCNQ2 | NM_172107.4 | chr20:g.63444715C>T | c.634G>A | p.(Asp212Asn) | Missense | AD | Heterozygous | Unknown | Unable to assess | | | Unlikely amenable |
| A144 | TSC1 | NM_000368.5 | chr9:g.132905731GCTGT>G | c.1843_1846del | p.(Thr615Profs*13) | Frameshift | AD | Heterozygous | LoF | Not eligible | | Possibly for wild type upregulation | Unlikely amenable |
| A145 | KCNQ2 | NM_172107.4 | chr20:g.63415086G>A | c.1342C>T | p.(Arg448*) | Nonsense | AD | Heterozygous | LoF | Not eligible | | Possibly for wild type upregulation | **Probably amenable** |
| A146 | SCN2A | NM_001040142.2 | chr2:g.165389334A>G | c.5528A>G | p.(Asp1843Gly) | Missense | AD | Heterozygous (possibly mosaic) | Unknown (likely GoF by phenotype) | Unable to assess | **Would be eligible for knockdown** | | Unlikely amenable |
| A147 | MTR | NM_000254.3 | chr1:g.236812761T>C | c.526T>C | p.(Tyr176His) | Missense | AR | Compound heterozygous | LoF | Not eligible | | | Unlikely amenable |
| A147 | MTR | NM_000254.3 | chr1:g.236891330G>A | c.3204+1G>A | p.? | Intronic SNV | AR | Compound heterozygous | LoF | Not eligible | | | Unlikely amenable |
| A148 | CSNK2B | NM_001320.7 | chr6:g.31669319G>A | c.368G>A | p.(Gly123Asp) | Missense | AD | Heterozygous | LoF | Not eligible | | | Unlikely amenable |
| A149 | IRF2BPL | NM_024496.4 | chr14:g.77027417G>A | c.376C>T | p.(Gln126*) | Nonsense | AD | Heterozygous | LoF | Not eligible | | Possibly for wild type upregulation | Unlikely amenable |
| A150 | TNRC6B | NM_001162501.2 | chr22:g.40265420GA>G | c.1192del | p.(Met398Trpfs*105) | Frameshift | AD | Heterozygous | LoF | Not eligible | | Possibly for wild type upregulation | Unlikely amenable |
| A151 | SCN1A | NM_001165963.4 | chr2:g.165991733G>A | c.5542C>T | p.(Gln1848*) | Nonsense | AD | Heterozygous (possibly mosaic) | LoF | **Eligible for WT upregulation** | | | Unlikely amenable |
| A152 | ATP7A | NM_000052.7 | chrX:g.78015858TG>T | c.2605del | p.(Val869*) | Nonsense | XLR | Hemizygous | LoF | Not eligible | | | Unlikely amenable |
| A153 | PNPLA6 | NM_001166114.2 | chr19:g.7550043C>T | c.1745C>T | p.(Pro582Leu) | Missense | AR | Homozygous | LoF | Not eligible | | | Unlikely amenable |
| A154 | PIGN | NM_176787.5 | chr18:g.62106885G>C | c.1675-4C>G | p.? | Intronic SNV | AR | Compound heterozygous | Unknown | Not eligible | | | Unlikely amenable |
| A154 | PIGN | NM_176787.5 | chr18:g.62072700T>TGGCA | c.2644_2645insTGCC | p.(Tyr882Leufs*7) | Frameshift | AR | Compound heterozygous | LoF | Not eligible | | | Unlikely amenable |
| A155 | SCN1A | NM_001165963.4 | chr2:g.165992278G>A | c.4997C>T | p.(Ser1666Phe) | Missense | AD | Heterozygous | Unknown (likely LoF by phenotype) | Unable to assess | **Would be eligible for WT upregulation** | | Unlikely amenable |
| A156 | PRRT2 | NM_145239.3 | chr16:g.29813694G>GC | c.649dup | p.(Arg217Profs*8) | Frameshift | AD | Heterozygous | LoF | Not eligible | | Possibly for wild type upregulation | Unlikely amenable |
| A157 | PRRT2 | NM_145239.3 | chr16:g.29813694G>GC | c.649dup | p.(Arg217Profs*8) | Frameshift | AD | Heterozygous | LoF | Not eligible | | Possibly for wild type upregulation | Unlikely amenable |
| A158 | PRRT2 | NM_145239.3 | chr16:g.29813694G>GC | c.649dup | p.(Arg217Profs*8) | Frameshift | AD | Heterozygous | LoF | Not eligible | | Possibly for wild type upregulation | Unlikely amenable |
| A159 | SCN1A | NM_001165963.4 | chr2:g.166043836T>C | c.1876A>G | p.(Ser626Gly) | Missense | AD | Heterozygous | Unknown (likely LoF by phenotype) | Unable to assess | **Would be eligible for WT upregulation** | | Unlikely amenable |
| A160 | PACS1 | NM_018026.3 | chr11:g.66211206C>T | c.607C>T | p.(Arg203Trp) | Missense | AD | Heterozygous | DN | **Eligible for knockdown** | | | Unlikely amenable |
